# Factors associated with the near miss of pregnant and postpartum women hospitalized by Covid-19

**DOI:** 10.1101/2023.05.10.23289809

**Authors:** Ana Carolina Schultz da Silva, Márcia Moroskoski, Natan Nascimento de Oliveira, Franciele Aline Machado de Brito, Marcela de Andrade Pereira Silva, Maria Aparecida Salci, Lígia Carreira, João Ricardo Nickenig Vissoci, Rosana Rosseto de Oliveira

**Author notes:** Corresponding author: João Ricardo Nickenig Vissoci.

## Abstract

**Objective:** To analyze the factors associated with maternal near miss due to Covid-19 in Brazil.

**Method:** cross-sectional study that assessed the cases of pregnant and postpartum women hospitalized with SARS due to Covid-19, aged between 10 and 49 years and from March 2020 to March 2022. Secondary data available in the Influenza Epidemiological Surveillance Information System (SIVEP-Flu) were used. Data were analyzed using SPSS statistical software, with univariate analysis followed by logistic regression.

**Results:** the significant independent factors associated with near miss in pregnant postpartum women due to Covid-19 were black/brown race, O2 saturation <95%, dyspnea, comorbidities, need for invasive and non-invasive ventilatory support.

**Conclusion:** the factors associated with near misses in pregnant and postpartum women with covid-19 reveal characteristics and risks inherent to the pathophysiology of the disease that contribute to worsening the condition.

## INTRODUCTION

In December 2019, the world was surprised by the emergence of a new virus in China, SARS-CoV-2. Already in March 2020, the World Health Organization (WHO) had declared it a pandemic due to its high transmissibility and the alarming number of cases and deaths in several countries and continents.^1^

Studies show an increased mortality rate from Covid-19 to the vulnerability of different population groups, with or without morbidity, and in different age groups.^2^ Among the most vulnerable groups are pregnant women, who are susceptible to severe infections due to physiological changes during pregnancy.^3^

In addition, although research has not shown evidence of vertical transmission of the virus, there is an increased prevalence of preterm birth, miscarriage, emergency cesarean section, intrauterine growth restriction, maternal death, maternal near misses, and other complications during pregnancy in women with Covid-19.^4^

In Brazil, the Covid-19 Observatory of the Oswaldo Cruz Foundation (Fiocruz) announced in 2021 that the country has the highest number of maternal deaths caused by the Covid-19 virus.^5^ Another study showed that about eight out of 10 pregnant and postpartum women who have become victims of the new coronavirus in the world are Brazilian.^6^ This rate could be even higher due to underreporting of cases and the fact that many women of reproductive age (10-49 years) have died.^7^ Reducing maternal mortality is a global challenge for health systems and has become even more challenging with the advent of Covid-19.^8^ The United Nations (UN), through the Sustainable Development Goals, agreed to the Brazilian reality of maternal mortality rate not exceeding 30 deaths per 100,000 live births by 2030.^9^ However, in 2021, this rate was 107 deaths per 100,000 births.^10^

The target set by the United Nations will only be possible through measures to improve the capacity to care for pregnant women before, during, and after childbirth.^8^ On the other hand, non-compliance will indicate the ineffectiveness of public policies aimed at women, since most cases are considered preventable deaths.^11^

In this sense, one of the strategies to reduce maternal mortality is to study the Near Miss Materno (NMM), which consists of analyzing cases in which the woman almost died due to a serious complication that occurred during pregnancy, childbirth, or the puerperium.^12^

It is believed that hospitalization of a pregnant or postpartum woman in intensive care for Covid-19 could be avoided through prevention and health promotion. Therefore, it is of paramount importance to study and train professionals working in these areas to recognize the factors associated with complications and thus avoid them.^13^

Therefore, this study aims to analyze the factors associated with maternal near misses due to Covid-19 in Brazil, considering the importance of the topic and the need to deepen the discussion and knowledge about this disease.

## METHOD

This is an observational, cross-sectional, and analytical study of notifications of pregnant and postpartum women hospitalized for Covid-19 in Brazil from March 2020 to March 2022. The study is based on the STROBE (Strengthening the Reporting of Observational Studies in Epidemiology) tool, which aims to strengthen the appropriate reporting of observational studies in epidemiology.^14^

The notifications analyzed correspond to the 27 states of the country, with an estimated population of 214.3 million inhabitants in 2022.^15^ The age range of 10 to 49 years was used because it is defined in Brazil as the group of women of childbearing age^16^, filtering also the cases of pregnant and postpartum women positive to Covid-19.

The notifications used were recorded in the public records of the Influenza Epidemiological Information System (SIVEP-Influenza) of the Ministry of Health (MS). SIVEP-Influenza is a surveillance program for the influenza virus in the country, from the sentinel surveillance network of the influenza syndrome. Due to the pandemic, the Health Surveillance Secretariat of the Ministry of Health (SVS/MS) adapted the system to include Covid-19 data.^17^ The data used in the study were extracted from the platform on April 15, 2021 using R software.

To achieve the objective of analyzing the factors associated with maternal near misses due to Covid-19, the dependent variable was the hospitalization of the pregnant and/or postpartum woman for Covid-19 in the ICU. Pregnant and postpartum women who died regardless of ICU or ward admission were excluded. Independent variables were grouped into sociodemographic characteristics (race, age, education, area of residence, and region); signs and symptoms (fever, sore throat, dyspnea, respiratory distress, saturation <95%, vomiting, diarrhea, and other symptoms); pre-existing conditions (comorbidity, cardiac, hematologic, neurologic, renal, hepatic, Down syndrome, asthma, diabetes, obesity, pneumonia, immunodeficiency); and clinical characteristics (obstetric population, ventilatory support, nosocomial infection, influenza vaccine, Covid-19 vaccine, antiviral).

The statistic used was logistic regression, which aims to construct a model from a set of observations that allows the prediction of the values taken by a categorical variable, in order to identify the factors that characterize each group of sick individuals compared to healthy individuals.^18^ Thus, in the univariate regression analysis, the variables with p <0.20 were selected.

The forward stepwise logistic regression model was then used, and the measures of association were the adjusted odds ratio (adjusted OR) with a 95% confidence interval (CI), taking into account the absence of multicollinearity in the model. Data were analyzed using the Statistical Package for the Social Sciences (SPSS, version 20.1, IBM, USA).

The study used secondary and public domain databases and did not require the approval of an ethics committee, according to the recommendations of Resolution No. 510/2016 of the National Health Council.^19^

## RESULTS

We analyzed 19,908 hospitalizations of pregnant and postpartum women with Covid-19 that occurred in Brazil between March 2020 and March 2022. Of these total hospitalizations, 16,358 (82.16%) were pregnant women and 3,550 (17.84%) were postpartum women. Considering the obstetric population, 4,132 (20.75%) experienced the situation of a near miss due to Covid-19, represented by ICU admission, and 15,776 (79.25%) were admitted to wards (Table 1).

**Table 1.** Univariate analysis of hospitalizations of pregnant and postpartum women with Covid-19, according to sociodemographic characteristics. Brazil, 2020-2022

|  | UTI |  | Infirmary |  | Total | OR | IC | p-value |
| --- | --- | --- | --- | --- | --- | --- | --- | --- |
|  | n (4132) | % | n (15776) | % |  |  |  |  |
| **Region** |  |  |  |  |  |  |  |  |
| South | 644 | 15,59 | 2796 | 17,72 | 3440 | 1,4 | 1,21-1,62 | <0,001 |
| Southeast | 1969 | 47,65 | 6449 | 40,88 | 8418 | 1,86 | 1,63-2,11 | <0,001 |
| Midwest | 397 | 9,61 | 1325 | 8,40 | 1722 | 1,82 | 1,55-2,15 | <0,001 |
| Northeast | 808 | 19,55 | 3297 | 20,90 | 4105 | 1,49 | 1,29-1,72 | <0,001 |
| North | 314 | 7,60 | 1909 | 12,10 | 2223 | - | - | - |
| **Age** |  |  |  |  |  |  |  |  |
| <20 | 259 | 6,27 | 1456 | 9,23 | 1715 | 0,73 | 0,634-0,838 | <0,001 |
| 20 to 34 | 2578 | 62,39 | 10564 | 66,96 | 13142 | - | - | - |
| ≥35 | 1295 | 31,34 | 3756 | 23,81 | 5051 | 1,41 | 1,309-1,525 | <0,001 |
| **Race/Color** |  |  |  |  |  |  |  |  |
| White | 1566 | 37,90 | 5685 | 36,04 | 7251 | - | - | - |
| Black/Brown | 1885 | 45,62 | 7619 | 48,29 | 9504 | 0,90 | 0,833-0,968 | 0,005 |
| Yellow | 28 | 0,68 | 113 | 0,72 | 141 | 0,90 | 0,593-1,366 | 0,619 |
| Indigenous | 12 | 0,29 | 138 | 0,87 | 150 | 0,32 | 0,175-0,571 | <0,001 |
| **Education** |  |  |  |  |  |  |  |  |
| Illiterate | 24 | 0,58 | 44 | 0,28 | 68 | 1,81 | 1,087-3,021 | 0,023 |
| Elementary | 414 | 10,02 | 1930 | 12,23 | 2344 | 0,71 | 0,609-0,835 | <0,001 |
| High School | 815 | 19,72 | 3761 | 23,84 | 4576 | 0,72 | 0,626-0,828 | <0,001 |
| College | 366 | 8,86 | 1216 | 7,71 | 1582 | - | - | - |
| **Zone** |  | 0,00 |  | 0,00 |  |  |  |  |
| Urban | 3391 | 82,07 | 13352 | 84,63 | 16743 | - | - | - |
| Rural | 216 | 5,23 | 1018 | 6,45 | 1234 | 0,83 | 0,718-0,972 | 0,02 |
| Peri-urban | 28 | 0,68 | 66 | 0,42 | 94 | 1,67 | 1,072-2,603 | 0,023 |

The sociodemographic aspects of pregnant and postpartum women with Covid-19 hospitalized in Brazil are shown in Table 1. The odds of ICU admission were higher in women aged 35 years and older (OR=1.41; CI=1.309-1.525), illiterate (OR=1.81; CI=1.087-3.021) and living in peri-urban areas (OR=1.67; CI=1.072-2.603). Among the Brazilian regions, the Southeast region had the highest odds ratio for the outcome (OR=1.86; CI=1.63-2.11) (Table 1).

Regarding the signs and symptoms of hospitalized pregnant and postpartum women with Covid-19 (Table 2), the results show that having had a fever (OR=1.47; CI=1.362-1.591), cough (OR=1.71; CI=1. 562-1.869), dyspnea (OR=4.57; CI=4.177-5.012), respiratory distress (OR=3.51; CI=3.231-3.810) and O2 saturation <95% (OR=5.62; CI=5.174-6.100) were factors associated with ICU admission. The sore throat was a protective factor for ICU admission (OR=0.77; CI=0.700-0.843).

**Table 2.**
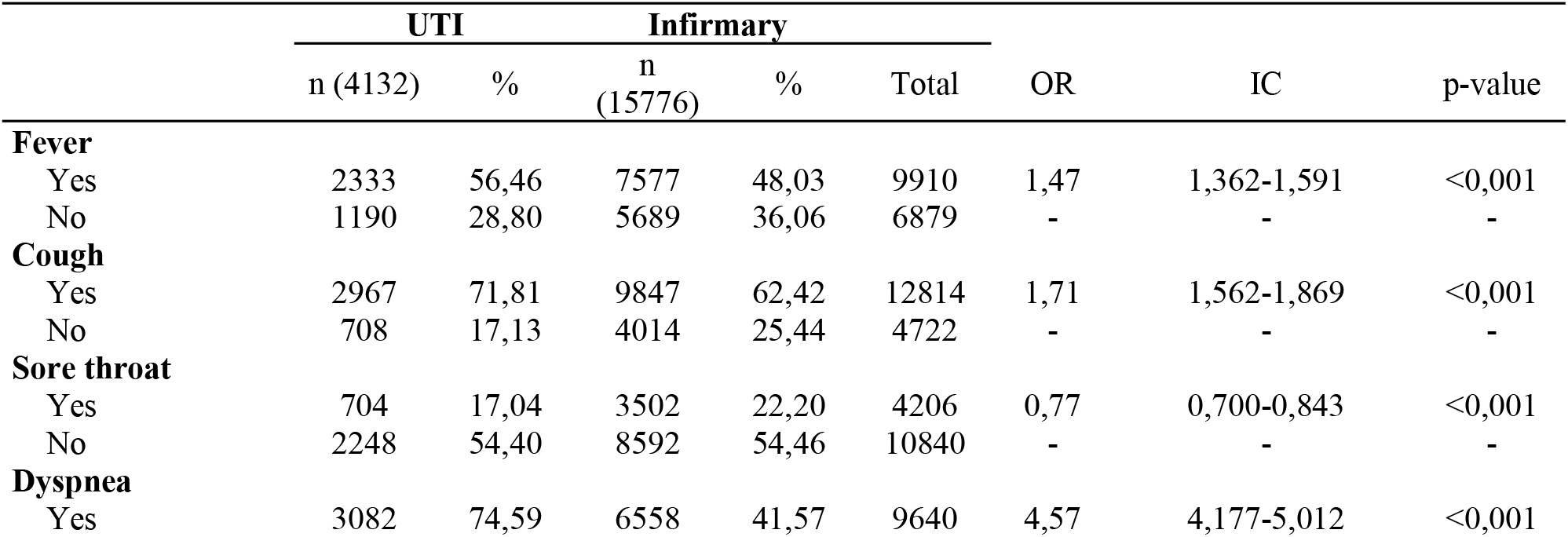
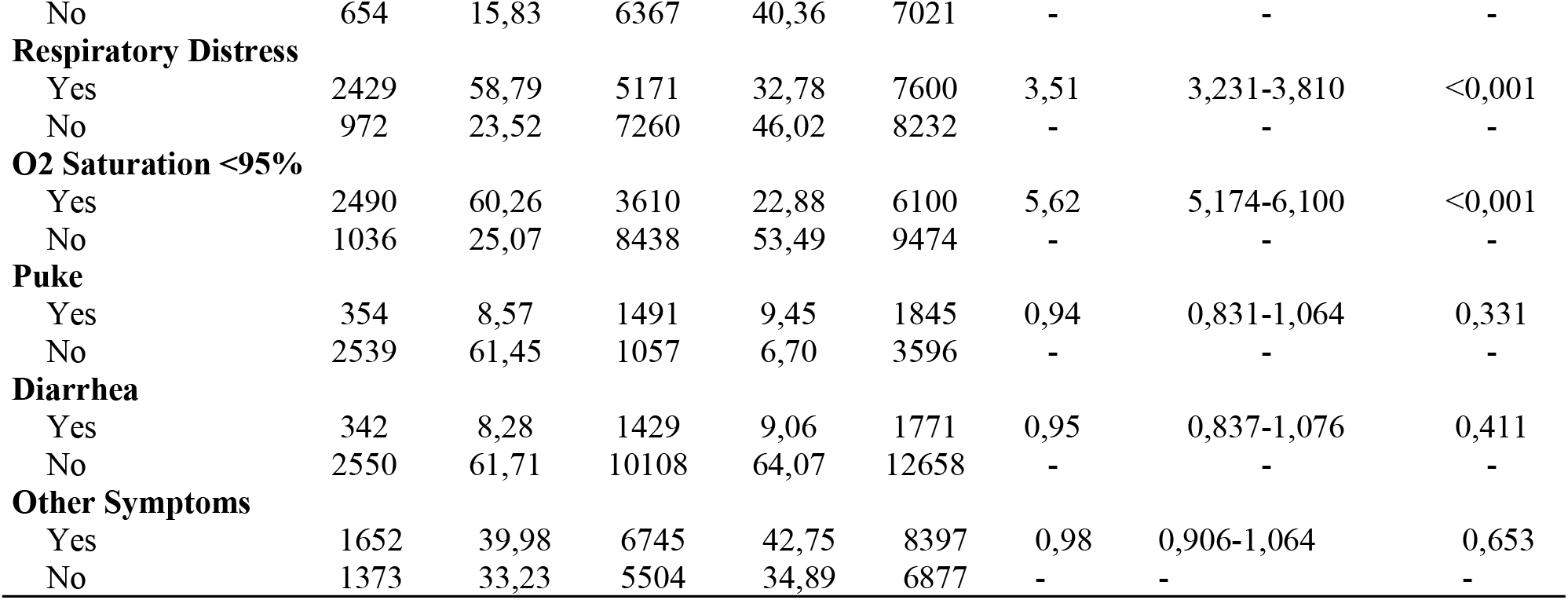
Univariate Analysis of Hospitalizations of Pregnant and Postpartum Women with Covid-19 by Signs and Symptoms Brazil, 2020-2022

Among the pre-existing comorbidities in pregnant and postpartum women with Covid-19, the diagnosis of heart disease (OR=1.49; CI=1.282-1.721), asthma (OR=1.45; CI=1.205-1.735), diabetes (OR=1.20; CI=1. 039-1.381), obesity (OR=2.37; CI=2.053-2.733), kidney disease (OR=1.69; CI=1.132-2.510) or the simple presence of comorbidity (OR=1.53; CI=1.423-1.639) were associated with a maternal near miss (Table 3).

**Table 3.** Univariate analysis of hospitalizations of pregnant and postpartum women with Covid-19 according to pre-existing comorbidities. Brazil, 2020-2022

|  | UTI |  | Infirmary |  | Total | OR | IC | p-value |
| --- | --- | --- | --- | --- | --- | --- | --- | --- |
|  | n (4132) | % | n (15776) | % |  |  |  |  |
| **Comorbidities** |  |  |  |  |  |  |  |  |
| Yes | 2604 | 63,02 | 8320 | 52,74 | 10924 | 1,53 | 1,423-1,639 | <0,001 |
| No | 1528 | 36,98 | 7456 | 47,26 | 8984 | - | - | - |
| **Cardiopathy** |  |  |  |  |  |  |  |  |
| Yes | 304 | 7,36 | 729 | 4,62 | 1033 | 1,49 | 1,282-1,721 | <0,001 |
| No | 1312 | 31,75 | 4674 | 29,63 | 5986 | - | - | - |
| **Haematological disease** |  |  |  |  |  |  |  |  |
| Yes | 28 | 0,68 | 65 | 0,41 | 93 | 1,46 | 0,935-2,286 | 0,096 |
| No | 1512 | 36,59 | 5132 | 32,53 | 6644 | - | - | - |
| **Down syndrome** |  |  |  |  |  |  |  |  |
| Yes | 7 | 0,17 | 23 | 0,15 | 30 | 1,03 | 0,440-2,397 | 0,952 |
| No | 1531 | 37,05 | 5164 | 32,73 | 6695 | - | - | - |
| **Liver disease** |  |  |  |  |  |  |  |  |
| Yes | 10 | 0,24 | 32 | 0,20 | 42 | 1,06 | 0,520-2,161 | 0,873 |
| No | 1516 | 36,69 | 5141 | 32,59 | 6657 | - | - | - |
| **Asthma** |  |  |  |  |  |  |  |  |
| Yes | 184 | 4,45 | 439 | 2,78 | 623 | 1,45 | 1,205-1,735 | <0,001 |
| No | 1406 | 34,03 | 4851 | 30,75 | 6257 | - | - | - |
| **Diabetes** |  |  |  |  |  |  |  |  |
| Yes | 314 | 7,60 | 895 | 5,67 | 1209 | 1,20 | 1,039-1,381 | 0,013 |
| No | 1332 | 32,24 | 4548 | 28,83 | 5880 | - | - | - |
| **Obesity** |  |  |  |  |  |  |  |  |
| Yes | 380 | 9,20 | 592 | 3,75 | 972 | 2,37 | 2,053-2,733 | <0,001 |
| No | 1268 | 30,69 | 4679 | 29,66 | 5947 | - | - | - |
| **Neurological disease** |  |  |  |  |  |  |  |  |
| Yes | 32 | 0,77 | 75 | 0,48 | 107 | 1,45 | 0,954-2,201 | 0,082 |
| No | 1504 | 36,40 | 5109 | 32,38 | 6613 | - | - | - |
| **Pneumopatias** |  |  |  |  |  |  |  |  |
| Yes | 29 | 0,70 | 75 | 0,48 | 104 | 1,31 | 0,852-2,024 | 0,217 |
| No | 1505 | 36,42 | 5112 | 32,40 | 6617 | - | - | - |
| **Immunodeficiency** |  |  |  |  |  |  |  |  |
| Yes | 34 | 0,82 | 123 | 0,78 | 157 | 0,94 | 0,639-1,378 | 0,747 |
| No | 1491 | 36,08 | 5063 | 32,09 | 6554 | - | - | - |
| **Kidney disease** |  |  |  |  |  |  |  |  |
| Yes | 37 | 0,90 | 75 | 0,48 | 112 | 1,69 | 1,132-2,510 | 0,010 |
| No | 1486 | 35,96 | 5078 | 32,19 | 6564 | - | - | - |

Regarding the analysis of clinical characteristics of pregnant and postpartum women hospitalized with Covid-19, it was associated with ICU admission being postpartum (OR=1.41; CI=1.299-1.538), having received invasive ventilatory support (OR=94.77; CI=79. 039-113.644), non-invasive ventilation (OR=5.53; CI=5.044-6.068), not having been vaccinated against influenza (OR=1.40; CI=1.243-1.585) and/or against Covid-19 (OR=1.71; CI=1.520-1.924) and having received antiviral treatment (OR=1.14; CI=1.003-1.290) (Table 4).

**Table 4.**
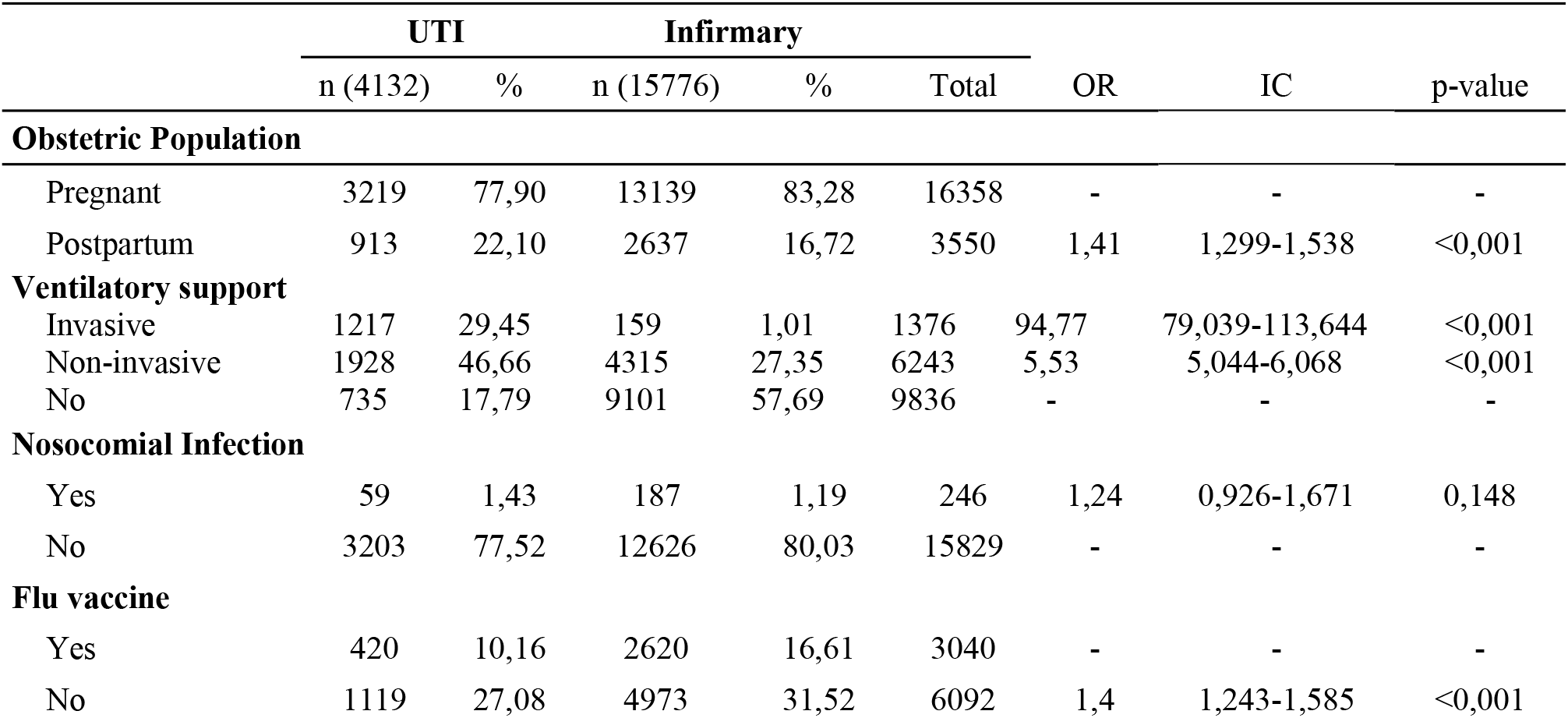
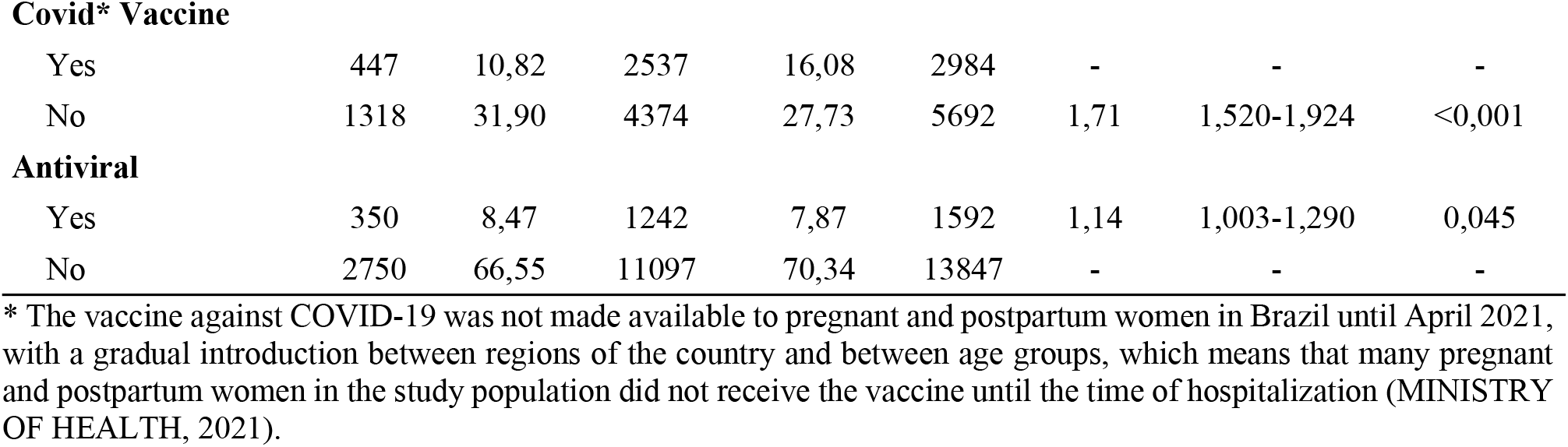
Univariate Analysis of Hospitalizations of Pregnant and Postpartum Women with Covid-19 by Clinical Characteristics Brazil, 2020-2022

|  | UTI |  | Infirmary |  | Total | OR | IC | p-value |
| --- | --- | --- | --- | --- | --- | --- | --- | --- |
|  | n (4132) | % | n (15776) | % |  |  |  |  |
| Obstetric Population |  |  |  |  |  |  |  |  |
| Pregnant | 3219 | 77,90 | 13139 | 83,28 | 16358 | - | - | - |
| Postpartum | 913 | 22,10 | 2637 | 16,72 | 3550 | 1,41 | 1,299-1,538 | <0,001 |
| Ventilatory support |  |  |  |  |  |  |  |  |
| Invasive | 1217 | 29,45 | 159 | 1,01 | 1376 | 94,77 | 79,039-113,644 | <0,001 |
| Non-invasive | 1928 | 46,66 | 4315 | 27,35 | 6243 | 5,53 | 5,044-6,068 | <0,001 |
| No | 735 | 17,79 | 9101 | 57,69 | 9836 | - | - | - |
| Nosocomial Infection |  |  |  |  |  |  |  |  |
| Yes | 59 | 1,43 | 187 | 1,19 | 246 | 1,24 | 0,926-1,671 | 0,148 |
| No | 3203 | 77,52 | 12626 | 80,03 | 15829 | - | - | - |
| Flu vaccine |  |  |  |  |  |  |  |  |
| Yes | 420 | 10,16 | 2620 | 16,61 | 3040 | - | - | - |
| No | 1119 | 27,08 | 4973 | 31,52 | 6092 | 1,4 | 1,243-1,585 | <0,001 |
| **Covid\* Vaccine** |  |  |  |  |  |  |  |  |
| Yes | 447 | 10,82 | 2537 | 16,08 | 2984 | - | - | - |
| No | 1318 | 31,90 | 4374 | 27,73 | 5692 | 1,71 | 1,520-1,924 | <0,001 |
| **Antiviral** |  |  |  |  |  |  |  |  |
| Yes | 350 | 8,47 | 1242 | 7,87 | 1592 | 1,14 | 1,003-1,290 | 0,045 |
| No | 2750 | 66,55 | 11097 | 70,34 | 13847 | - | - | - |

\* The vaccine against COVID-19 was not made available to pregnant and postpartum women in Brazil until April 2021, with a gradual introduction between regions of the country and between age groups, which means that many pregnant and postpartum women in the study population did not receive the vaccine until the time of hospitalization (MINISTRY OF HEALTH, 2021).

In logistic regression analysis, the independent significant factors associated with a near miss in pregnant and postpartum women with Covid-19 were being black/brown (ORaj=0.83; CI=0.720-0.966), having O2 saturation <95% (ORaj=1.92; CI=1.609-2.299), presenting dyspnea (ORaj=1.47; CI=1.216-1.778), having comorbidities (ORaj=1. 49; CI=1.282-1.737), requiring invasive mechanical ventilation (ORaj=44.99; CI=33.099-61.167), requiring non-invasive mechanical ventilation (ORaj=3.32; CI=2.754-3.994), and not having been vaccinated against influenza (ORaj=1.18; CI=1.006-1.390), with all these factors controlled by yellow and indigenous race/color (Table 5).

**Table 5.** Final multiple logistic regression model of factors associated with outcome in hospitalized pregnant and postpartum women with Covid-19, according to ICU admission. Brazil, 2020-2022

|  | ORaj* | IC(95%) | p |
| --- | --- | --- | --- |
| **Maternal characteristics** |  |  |  |
| Race/black/brown color | 0,83 | 0,720-0,966 | 0,016 |
| **Signs and symptoms** |  |  |  |
| O2 Saturation <95% | 1,92 | 1,609-2,299 | <0,001 |
| Dyspnea | 1,47 | 1,216-1,778 | <0,001 |
| **Pre-existing comorbidities** |  |  |  |
| Comorbidity present | 1,49 | 1,282-1,737 | <0,001 |
| **Clinical features** |  |  |  |
| Invasive ventilatory support | 44,99 | 33,099-61,167 | <0,001 |
| Noninvasive ventilatory support | 3,32 | 2,754-3,994 | <0,001 |
| Not having a flu vaccine | 1,18 | 1,006-1,390 | 0,042 |

\* Adjusted for yellow and indigenous race/color.

## DISCUSSION

This study was able to identify the characteristics of women who developed a maternal near miss due to Covid-19. Factors associated with maternal near misses were black/brown color, history of comorbidities, O2 saturation <95%, dyspnea, need for invasive and non-invasive ventilatory support, and no influenza vaccination.

Regarding the socio-demographic profile, black and brown women were predominant, demonstrating that they are more susceptible to Covid-19 infection. However, the proportion of black and brown women admitted to intensive care was lower, which may be related to their lack of access to health services and technologies. The smaller number of black and brown pregnant and postpartum women admitted to ICU beds compared with those admitted to wards confounded the odds ratio, and black/brown race/color could be confused with a protective factor. However, a large body of literature shows the opposite, with whites being the most hospitalized population and black/browns having higher mortality rates.^20^

In this sense, the historical difficulties of this population group in accessing the health system are notorious, as evidenced by unfavorable indicators such as maternal mortality, since they represent approximately 65% of the total maternal deaths in the country.^21^

Another important aspect is the social inequality between whites and blacks/brown. A survey conducted during the pandemic showed that the per capita household income of the white Brazilian population is much higher. In July 2020, the per capita income of whites was R$ 1,639.00, while that of the black and brown population was R$ 971.00.^22^ Therefore, social inequalities deserve attention due to their strong relationship with situational health indicators.

In terms of signs and symptoms, there was a predominance of O2 saturation <95% and dyspnea. Physiologically and anatomically, pregnant women undergo pregnancy-related changes that increase the risk of complications and death from Covid-19. Pulmonary physiology undergoes hormonal and functional changes that make it less tolerant to hypoxia. In early pregnancy, progesterone levels act on the brainstem to increase respiratory rate and tidal volume, and chest wall compliance and airway resistance decrease. In the last trimester, the uterus constricts the diaphragm, reducing total lung capacity.^23^

In addition to these respiratory changes, there are important immunologic changes that place pregnant and postpartum women at risk for developing more severe respiratory infections, which can lead to hypoxemia, which in combination with pulmonary infection can lead to vasoconstriction and worsening of the condition.^23^

Dyspnea is very common in Covid-19, reported in 81% of hospitalized patients. The presence of this symptom is related to the severity of the disease, being less common in the early phase.^24^ When present, dyspnea occurs because the disease uses angiotensin-converting enzyme 2 (ACE 2) as a receptor, which is widely distributed in pulmonary, intestinal, renal, and vascular tissues. As a result, cytokines are released that damage lung, kidney, and heart tissue and worsen organ system function.^25^ Among pre-existing comorbidities, cardiac disease, hematologic disease, diabetes, asthma, neurologic disease, renal disease, pulmonary disease, and obesity are considered risk factors for adverse outcomes in coronavirus-infected patients.^26-27^

In this regard, the present study found that most pregnant and postpartum women had comorbidities. A review of 77 studies concluded that advanced maternal age, high body mass index, and previous comorbidities were the most relevant risk factors in pregnant women who developed the most severe form of the disease and required intensive care. These findings suggest a risk profile for adverse maternal outcomes such as ICU admission and clinical deterioration, which may progress to death.^28^

Among the most relevant clinical characteristics, the use of ventilatory support (invasive and noninvasive) stands out. Studies conducted in Brazil showed that 20% of pregnant and postpartum women with the severe acute respiratory syndrome (SARS) required ICU admission, 22% required noninvasive ventilation, and 10 to 25% required invasive ventilation.^29-30^

In light of Covid-19 and SARS, ventilatory support has become a common requirement in the management of affected patients. In several contexts, the use of invasive techniques for oxygenation was higher than that of non-invasive techniques, with a significant increase in 2021, due to the prolonged and fatal nature of the disease, indicating the intensification of care to reverse the severe condition.^31^

Regarding vaccination against influenza (H1N1), an association has been observed with hospitalizations for Covid-19. In this sense, unvaccinated patients had a higher risk of ICU admission, confirming studies showing that vaccination reduces the number of positive tests for Covid-19 and improves clinical outcomes.^32^

Another study suggests that influenza vaccination is associated with a lower risk of cardiovascular events and reduces the likelihood of overlapping secondary infections, as patients with Covid-19 can acquire the influenza virus at the same time.^33^

Although some international studies have shown high morbidity and mortality in pregnant and postpartum women with the severe form of COVID-19, the clinical course of the disease in pregnant women from other countries has not shown a significant increase in the rate of outcomes with worse prognosis compared to the general population and the Brazilian reality.^34-35^

As a result, WHO, together with the American College of Gynecology and Obstetrics, observed an increase in weekly deaths among pregnant and postpartum women and included this group as a priority in the country’s vaccination campaigns as an important preventive measure, given that the effectiveness of the vaccine is comparable to that of nonpregnant women.^36^ In light of these findings and the pandemic scenario, it is essential that health services promote the vaccination of the obstetric population, which has been shown to have an impact on the development of severe cases, particularly respiratory disease.

Given these findings and the pandemic scenario, health services must promote the vaccination of the obstetric population, which has been shown to have a positive impact on the development of severe cases, particularly those with respiratory failure requiring ventilatory support and invasive ventilation, reaffirming the importance of vaccination against Covid-19.

It is also necessary to implement new protocols and preventive measures aimed at combating and preventing new transmissions of the virus, culminating in new configurations and adaptations of assistance to pregnant and postpartum women. In addition, policies that address racial inequalities are essential to contribute to the improvement of care, which must comply with the principles of the Single Health System (SUS) in terms of universality, inclusiveness, and equity.

It is worth mentioning that the present study has some limitations, mainly related to the use of secondary data, which are subject to underreporting or completion problems. However, the health information systems of the Brazilian Ministry of Health are of great value, as they allow the epidemiological study of diseases at the national level, which helps in the planning of health policies and programs. In addition, during the Covid-19 pandemic, SIVEP-Influenza was the main source of data on Covid-19 hospitalizations in the country.

## CONCLUSION

ICU admissions of pregnant and postpartum women with Covid-19, considered in this study as maternal near misses, were associated with black and brown race/color, with O2 saturation <95%, with dyspnea, with comorbidities, with a need for invasive or non-invasive ventilatory support, and with lack of influenza vaccination.

In this sense, recognition of the factors involved in maternal near misses can help to direct actions to qualify the care of women in pregnancy and the puerperium. In addition, it allows us to identify which women are more likely to have a fatal outcome, thus allowing early intervention.

## Data Availability

The data underlying the results presented in the study are available from https://opendatasus.saude.gov.br/

https://datasus.saude.gov.br/

## ACKNOWLEDGEMENTS

We would like to thank the Universidade Estadual de Maringá for their support in carrying out the research and the Centro Universitário Uningá and Duke University for the partnership.

## REFERENCES

1. Organização Mundial da Saúde (OMS). OMS declara pandemia do novo Coronavírus. Universidade Aberta do Sus (UNA-SUS). 11 de março de 2020 [Cited 2022 jan 23]. Available from: https://www.unasus.gov.br/noticia/organizacao-mundial-de-saude-declara-pandemia-de-coronavirus.

2. Zaigham M, Andersson O. (2020). Resultados maternos e perinatais com COVID-19: Uma revisão sistemática de 108 gestações. Acta Obstet Gynecol Scand. Available from: https://doi.org/10.1111/aogs.13867.

3. Yan J, Guo J, Fan C, et al. (2020). Coronavirus disease 2019 in pregnant women: a report based on 116 cases. Am J Obstet Gynecol, Available from: https://pubmed.ncbi.nlm.nih.gov/32335053/

4. Fernandes JN, Rezende MCD, Otsubo BYV, et al. (2022). Correlation between COVID-19 and gestational complications: a systematic review. Brazilian Journal of Health Review, v5, n 2, 6405–6411 [Cited 2023 apr 11]. Available from: https://doi.org/10.34119/bjhrv5n2-213

5. Fundação Oswaldo Cruz (Fiocruz). (2021). Boletim Extraordinário, 25 de junho de 2021. [Cited 2022 jan 24]. Available from: https://agencia.fiocruz.br/sites/agencia.fiocruz.br/files/u34/boletim_extraordinario_2021-junho-23-parte2-pags09-17.pdf

6. Bastos, LS, Niquini RP, Lana RM, et al. (2020) COVID-19 and hospitalizations for SARI in Brazil: a comparison up to the 12th epidemiological week of 2020. Cad Saúde Pública. Available from: https://www.scielo.br/j/csp/a/KQxzHZdFHcPx5CftPXZKwgs/?format=pdf&lang=en

7. Almeida JP, Santana VS, Santos KM, et al. (2021). Internações por SRAG e óbitos por Covid-19 em gestantes brasileiras: uma análise da triste realidade. Brazilian Journal of Health Review. Available from: https://www.brazilianjournals.com/index.php/BJHR/article/view/31570

8. Motta CT (2021). Subsidies for the compliance of SDG 3.1 of the 2030 Agenda: an analysis of maternal mortality in Brazil from 1996 to 2018. M. Sc. Dissertação, Escola Nacional de Saúde Pública Sérgio Arouca. Available from: https://www.arca.fiocruz.br/bitstream/handle/icict/48404/caio_tavares_motta_ensp_mest_2021.pdf;jsessionid=8396E5143894CDCB6684F3F1C8D35951?sequence=2

9. Instituto de Pesquisa Econômica Aplicada (IPEA). ODS – Metas Nacionais dos Objetivos de Desenvolvimento Sustentável. 2018. [Cited 2022 jan 24]. Available from: https://repositorio.ipea.gov.br/bitstream/11058/8855/1/Agenda_2030_ods_metas_nac_dos_obj_de_desenv_susten_propos_de_adequa.pdf

10. Observatório Obstétrico Brasileiro (OOBr). Óbitos de Gestantes e Puérperas. 2022. [Cited 2022 jan 24]. Available from: https://observatorioobstetrico.shinyapps.io/obitos-grav-puerp.

11. Souza ASR, Amorim MMR. (2021). Maternal mortality by Covid-19 in Brazil. Rev Bras Saúde Matern Infant. Available from: https://www.scielo.br/j/rbsmi/a/R7MkrnCgdmyMpBcL7x77QZd/?format=pdf&lang=en

12. Organização Mundial da Saúde (OMS). Avaliação da qualidade do cuidado nas complicações graves da gestação: A abordagem do near miss da OMS para a saúde materna. 2011. [Cited 2022 jan 27]. Available from: https://www.paho.org/clap/dmdocuments/CLAP-Trad05pt.pdf

13. Lima R. (2021). Direito sexuais e direitos reprodutivos das mulheres. Escola de Assistência Jurídica da Defensoria Pública. [Cited 2022 aug 18]. Available from: Cartilha DireitoSexuais e Reprodutivos Das Mulheres | PDF (http://scribd.com)

14. Malta M, Cardoso LO, Bastos FI et al. (2010) STROBE initiative: guidelines on reporting observational studies. Rev Saude Publica, v. 44, n. 3, p. 559–565. DOI: OBJ

15. Instituto Brasileiro de Geografia e Estatística (IBGE). (2022). Projeção da população do Brasil e das Unidades da Federação. [Cited 2022 jun 21]. Available from: https://ibge.gov.br/apps/populacao/projecao/

16. Brasil. (2008). Ministério da Saúde. Portaria nº 1,119, de 5 de junho de 2008. Regulamenta a vigilância de óbitos maternos. [Cited 2022 jun 21]. Available from: https://bvsms.saude.gov.br/bvs/saudelegis/gm/2008/prt1119_05_06_2008.html

17. Bahia. Secretária da Saúde do Estado da Bahia. Guia Rápido SIVEP – Gripe. 2020. [Cited 2022 jun 14]. Available from: https://docs.bvsalud.org/biblioref/2021/03/1147534/guia-rapido-sivep-gripe_agosto-2020.pdf

18. Garson GD. (2014). Logistic Regression: Binary & Multinomial [Cited 2022 aug 21]. Available from: file:///C:/Users/marci/Downloads/Logistic%20Regression_%20Binary%20and%20Multinomial%20(%20PDFDrive%20).pdf

19. Brasil. (2016). Resolução nº 510, de 7 de abril de 2016. [Cited 2022 jun 21]. Available from: https://conselho.saude.gov.br/resolucoes/2016/Reso510.pdf

20. Zeiser FA, Donida B, Costa CA, et al. (2022) First and second COVID-19 waves in Brazil: A cross-sectional study of patients’ characteristics related to hospitalization and in-hospital mortality. The Lancet Regional Health - Americas, v. 6, n. November, p. 1–16, 2022. DOI: https://doi.org/10.1016/j.lana.2021.100107

21. Lima LCBB. (2021). A ocorrência de falhas assistenciais durante o trabalho de parto: uma revisão integrativa. Trabalho de Conclusão de Curso. Pontifícia Universidade Católica de Goiás. [Cited 2023 mar 07]. Available from: https://repositorio.pucgoias.edu.br/jspui/handle/123456789/2510?mode=full

22. Instituto de Pesquisa Econômica e Aplicada (IPEA). (2021). A pandemia de Covid-19 e a desigualdade racial de renda. Boletim de Análise Político-Institucional, n. 26, março 2021. [Cited 2023 mar 08]. Available from: https://repositorio.ipea.gov.br/bitstream/11058/10519/1/BAPI_26_DesRacial.pdf

23. Czeresnia RM, Trad ATA, Britto ISW, et al. (2020). Sars-Cov2 and Pregnancy: a review of the facts. Rev Bras Ginecol Obstet,v42, 9, 562–568. [Cited 2022 aug 21]. Available from: https://doi.org/10.1055/s-0040-1715137

24. Nascimento IJB, Groote TC, O’Mathúna DP, et al. (2020). Clinical, laboratory and radiological chacrateristics and outcomes of nves coronavírus (SARS-CoV-2) infection in humans: A systematic review and series of meta-analýses. PLos One. [Cited 2022 aug 22]. Available from: 10.1371/journal.pone.0239235

25. Hoffmann M, Kleine-Weber H, Schoeder S, et al. (2020) SARS-CoV-2 cell entry depends on ACE2 and TMPRSS2 and is blocked by a clinically proven protease inhibitor. Cell, v 181, 271–280. [Cited 2022 Aug 22]. Available from: https://doi.org/10.1016/j.cell.2020.02.052

26. Elshafeey F, Magdi R, Hindi N, et al. (2020). Asistematic scoping review of Covid-19 during pregnancy and childbirth. International Journal of Gynecology & Obstetrics, v1, 2. [Cited 2022 Aug 21]. Available from: https://doi.org/10.1002/ijgo.13182

27. Jering KS, Claggett BL, Cunningham JW, et al. (2021). Clinical characteristics and outcomes of hospitalized women giving birth and without Covid-19. JAMA Intern Med, 181, v 5, 714–717. [Cited 2022 aug 21]. Available from: https://doi.org/10.1001/jamainternmed.2020.9241

28. Allotey J, Fernandez S, Bonet M, et al. (2020). Clinical manifestations, risk factors, and maternal and parinatal outcones of coronavírus diasease 2019 in pregnancy: living systematic review meta-analysis. The BMJ, 370, m3320. [Cited 2022 aug 22]. Available from: Available from: https://doi.org/10.1136/bmj.m3320

29. Godoi APN, Bernardes GCS, Almeida NA, et al. (2021) Severe Acute Respiratory Syndrome by COVID-19 in pregnant and postpartum women. Rev Bra Saúde Matern Infant, v 21, n 2, 5461–5469. [Cited 2022 aug 22]. Available from: https://doi.org/10.1590/1806-9304202100S200008

30. Leal LF, Merckx J, Fell DB, et al. (2021). Characteristics and outcomes of pregnant woman with SARS-CoV-2 infection and other severe acute respiratory infections (SARI) in Brazil from January to November 2020. Braz J Infect Dis. [Cited 2022 aug 24]. Available from: 10.1016/j.bjid.2021.101620

31. Takemoto MLS, Menezes MO, Andreucci CB, et al. (2020). The tragedy of COVID-19 in Brazil: 124 maternal deaths and counting. Int J Gynaecol Obstet, v 151, n 1, 154–156. [Cited 2022 aug 22]. Available from: Available from: https://doi.org/10.1002/ijgo.13300

32. Conlon A, Ashur C, Washer L. et al. (2021). Impacto f the influenza vaccine on Covid-19 infection rates and severity. American Journal of Infection Control, v 49, 694–700. [Cited 2022 aug 22]. Available from: https://doi.org/10.1016/j.ajic.2021.02.012

33. Behrouzi B, Campoverde MVA, Liang K, et al. (2020). Influenza Vaccination to Reduce Cardiovascular Morbidity and Mortality in Patients With COVID-19. Journal of the American College of Cardiology, v. 76, n. 15, p. 1–19, 2020. DOI: 10.1016/j.jacc.2020.08.028

34. Dashraath P, Wong JLJ, Lim MXK, et al. (2020). Coronavirus Diasease 2019 (COVID-19) pandemic and pregnancy. Am J Obstet Gynecol, v 222, n 6, 521–531. [Cited 2022 aug 18]. Available from: https://doi.org/10.106/j.ajog.2020.03.021

35. Pacagnella RCP, Nakamura-Pereira M, Gomes-Sponholz F, et al. (2018). Maternal mortality in Brazil: Proposals and strategies for its reduction. Rev Bras Ginecol Obstet, v 40, 501–506). [Cited 2022 aug 18]. Available from: https://doi.org/10.1055/s-0038-1672181

36. Rodrigues FO, Antunes Neto A, Silva RG, et al. (2021). Desfechos maternos da COVID-19 e atualizações sobe a vacinação em gestantes e puérperas. Revista Brazilian Journal of Development, v 7, n 6, 57232–57247. [Cited 2021 jan 27]. Available from: https://doi.org/10.34117/bjdv7n6-227

